# Artificial intelligence for automated thoracic aorta diameter measurement using different computed tomography protocols

**DOI:** 10.1101/2022.12.29.22284036

**Authors:** Maria Fernanda Portugal, Lucas Lembrança Pinheiro, Henrique Min Ho Lee, Henrique Cursino Vieira, Lariza Laura de Oliveira, Matheus del Valle, Newton Shydeo Brandão Miyoshi, Livia Oliveira-Ciabati, Ronaldo Barone, Gilberto Szarf, Nelson Wolosker

## Abstract

**Introduction:** Thoracic aortic aneurysm diameter determination is paramount for the decision-making process regarding surgical management. Studies focusing in asymptomatic patients have determined prevalence of 0.16 to 0.36% of TAAs in imaging studies. Several groups have proposed automated aortic measurement tools as propaedeutic and therapeutic instruments. In this study we developed and tested an automatic 3-dimensional (3D) segmentation method for the thoracic aorta, applicable on computed tomography angiography (CTA) acquired using low-dose and standard dose protocol, with and without contrast enhancement; and to accurately calculate the 3D diameter information of the arterial segments.

**Methods:** a retrospective cohort of all CT scans acquired in our service between 2016 and 2021 led to the selection of 587 CT exams including low and standard-dose radiation, with and without contrast enhancement. 527 exams were used for neural network training of an algorithm capable of aptly measuring the aortic diameters, using manual measurements performed by three medical specialists as a baseline. Sixty exams were used for validation. The algorithm was developed both for use with the support of PyRadiomics and for a self-made approach.

**Results:** Aortic measurement using the algorithm supported by PyRadiomics resulted in mean absolute error values under 2mm. For the self-made approach, mean absolute error values were under 5mm.

**Conclusion:** This study presents an effective automated solution for thoracic aortic measurement with good results in sets of standard or low-radiation exams, as well as those acquired with or without contrast enhancement; presenting a possibility for an auxiliary tool for automation of the process of measuring the diameter of the thoracic aorta.

## 1 Introduction

Aneurysms of the thoracic aorta (TAA) represent one third of hospital admissions from aortic diseases in the United States^1^. Although they fall into the classical definition aneurysms –arterial dilations of at least 50% the adjacent healthy segments^2,3^, aneurysms in the thoracic aorta are associated with a natural history vastly diverging from that described for their more frequent abdominal counterparts: they present slower growth and higher association to congenital and degenerative diseases^4^.

Several studies have attempted to assess the incidence of TAAs^5^. However, the key point for disease determination is dependent upon the average aortic size in a given population, which is broadly variable^1,6,7^. TAA diameter determination is also paramount for the decision-making process regarding surgical management^8^, with current guidelines proposing surgical treatment for dilations over 4cm in the ascending aorta and over 6cm in the descending segment, though these may vary according to comorbidities^1^.

Although screening strategies have been deemed effective for reducing mortality of abdominal aortic aneurysms^9^, in the thoracic segment, general assessment of all asymptomatic patient is not recommended by the American Heart Association does not recommend, unless they present clear risk factors, such as collagen-specific diseases or first-degree relative family history^1^. Nevertheless, studies focusing in asymptomatic patients have determined prevalence of 0.16 to 0.36% of TAAs in imaging studies^10–12^. On the other hand, annual imaging screenings for lung-cancer are recommended, using low-dose chest computed tomography (CT) scans. This recommendation applies for patients over 50 years of age and with a present of past history of smoking^13^ – both risk factors for aortic aneurysms^1^.

Several groups have proposed automated aortic measurement tools as propaedeutic and therapeutic instruments^14–16^. In particular, convolutional neural networks (CNNs) have achieved state-of-the-art results on a wide range of anatomical structures^16^. The present study aimed to develop and test an automatic 3-dimensional (3D) segmentation method for the thoracic aorta, applicable on computed tomography angiography (CTA) acquired using low-dose and standard dose protocol, with and without contrast enhancement; and to accurately calculate the 3D diameter information of the arterial segments, thus facilitating the estimation of aortic size in high-risk patients subjected to screening for lung cancer. This automatic method was trained and validated against manual measurements performed by three specialists (one Radiologist and two Vascular Surgeons) in 587 exams.

## 2 Methods

This study is a retrospective cohort of all CT scans acquired between 2016 and 2021, from the imaging dataset of our institution. This study was approved by the Ethics Committee under protocol number 44951021.8.0000.0071, through the report number 4.710.910 of the 13^th^ of May, 2021.

This study was funded under portents of the Law 8.248, of the 23^rd^ of October of 1991.

### 2.1 Data collection

CT studies were performed following institutional protocol for thoracic exams and the exam indication, using collimation up to 1.3 mm. For contrasted exams, iodine-based intravenous contrast medium (1–2 mL/kg of body weight) was delivered using a power injector, with variable injection rates (2.0–5.0 mL/s). Slice thickness of images was of up to 1.5 mm. Devices from different vendors were included (Siemens, General Electric and Canon).

Acquisitions were included if they did not present artefacts due to movement or presence of metal devices. All exams with known previously treated aneurysms, presence of endografts, active aortic dissections and valve grafts were excluded.

All data was prospectively entered into a dedicated anonymized database. After this step, the population characteristics were described by age, biological sex, weight, height, and the exam direction (head to feet or the opposite).

### 2.2 Manual image segmentation

The selected exams underwent a segmentation process by three expert doctors in the field: two vascular surgeons and one radiologist. A manual, slice-by-slice segmentation of each exam was done using the 3D Slicer software^17^, a free open-source software distributed under a BSD license style, using gold-standard segmentation method “thresholding”.

The totality of the thoracic aorta was highlighted, including the entire arterial wall visible in each slice, from just above the aortic valve to just above the diaphragm. Additionally, a centerline was marked using as reference points the center of aortic valve, the emergency of the brachiocephalic trunk, the emergency of the left subclavian artery and the diaphragmatic hiatus.

In all cases in which identification of the aortic valve was not possible, an arbitrary point 2.5cm proximal to the brachiocephalic trunk was considered as the aortic origin. In all cases in which identification of the diaphragmatic hiatus was not possible, the diaphragmatic cupula was considered as the inferior limit of the thoracic aorta.

A sample of exams was segmented in triplicate to be evaluated. These exams will be unified through the voting method and added to the other exams. The voting method consists of verifying whether at least two markings register at the same location, if not, the location is discarded.

### 2.3 Manual image segmentation between-comparison

The annotation of the aorta evaluation was performed by the three physicians and these measurements were compared using the Dice score coefficient metric. This metric aims to calculate the area overlap using the number of pixels in the image. A higher score coefficient indicates more overlapping images, and a more similar marking with regard to the determined gold-standard.

### 2.4 Pre-processing

To standardize the dataset, the patients’ positions were changed from FFS (feet first supine) to HFS (head first supine) and next step the intensity was adjusted by conversion of Hounsfield unit.

The images were also cropped to exclude regions below the diaphragm, when present. Resampling steps were applied, and isomorphic resolutions were kept preserving the voxel dimensions. The “nearest-neighbour” approach was used for resampling and reshaping, in order to preserve image-binary segmentation relation. Finally, the window level and width were adjusted to get images of the same size.

### 2.5 Neural Network training

The dataset was divided into training and validation datasets. For the training dataset, 90% of scans with their corresponding ground truth segmentations were randomly selected and used for neural network training (training dataset). For the validation dataset, the remaining 10% were used. Both sets included all exam types, as homogeneously distributed as possible.

Models were trained using a 4-fold cross-validation. The models’ performance was evaluated by use of the loss function values, as well as the Dice Score Coefficient (DSC). The architectures DeepAAA^18^ and DeepVox were the basis for Convolution Neural Networking (CNN) training. The DeepVox architecture was developed by our team specifically for this project.

The model conception was based on the Vox2Vox, a Conditional Generative Adversary Networking (cGAN)^19^, with variable z from DeepAAA. Other changes were added to increase performance, such as adding a VGG-11 based model to the discriminator. The losses were set like the Vox2Vox, except that hybrid focal loss was used instead of dice loss.

The deep learning models were trained using cross-validation and 100 epochs per fold. Images were compressed in the z-axis with a fixed voxel depth of 3 mm, instead of forcing a z shape of 128, while the x and y axes were downsampled by resampling and cropping, not reshaping, granting uniformity for all the exams, in all dimensions. The variable z-axis enables the model to receive both thoracic and thoracoabdominal exams, identifying where the thoracic aorta ends and not proceeding with the segmentation to the abdominal region.

### 2.6 Thoracic aorta diameter measurements

To assess the created automatic measurement system outcomes, four different methods of measuring the thoracic aorta were used. Those methods are: (i) manual measuring, (ii) semi-automatic measuring, (iii) automatic measuring based on a made-to-measure algorithm and (iv) automatic measuring based on PyRadiomics^20^.

#### 2.6.1 Manual measuring

The first method is manual measurement, the diameters of specific positions and the length of the ascending and descending aorta were measured manually using the Horus® software, an open-source tool developed by bq, and released to the community under GPLv2.

#### 2.6.2 Semi-automatic measuring

Images were segmented using the 3DSlicer, for semi-automatic aortic measurement. Diameters were measured using the VMTKSlicer Module, an add-on open source for 3DSlicer. This extension was also used to build a centerline to measure the thoracic aorta’s diameter. To manually construct the centerline was required for the user to centralize the starting and ending positions to serve as centerline’s reference points along the segmented aorta.

#### 2.6.3. Automatic measuring

Automatic measures of the aortic diameter were done by two techniques: PyRadiomics 3.0.1 and a made-to-measure algorithm (Supplementary Figure 1). The first approach uses an open-source python package to extract Radiomics features from medical imaging to obtain the diameter, whilst also calculating the smallest axis length. In the second method, an algorithm was developed to compare the greatest segment of the aorta with the regions declared healthy. The algorithm took the patient mask as input and the output was a list of average aortic diameter automatically measured. This algorithm obtained a centerline based on the skeletonizing method. The occasional errors created by the skeletonizing method were corrected using a graph-based pruning function, and then the algorithm determined each point of the centerline. A normalized vector x was calculated using the current point and the next point in the centerline. As a next step, the P orthogonal plane to ||x|| was calculated as well as the intersection of P with the aorta masks. Then, the Euclidian distance was calculated between the current point and all other points in the intersection. The pseudo-code of the algorithm is presented in (Supplementary Figure 1).

### 2.7 Aortic diameter comparison and statistical analysis

The mean size, standard deviation, and maximum value obtained from each measurement method of the thoracic aorta for all exams were extracted. The mean absolute error metric was used to compare the mean size between each measurement method. This metric consists of calculating the difference between all measurements from different ways for each patient and then averaging the absolute value of errors.

The dataset composition, according to exam type by modality and aneurysm presence, is shown in Figure 1.

**Figure 1:**
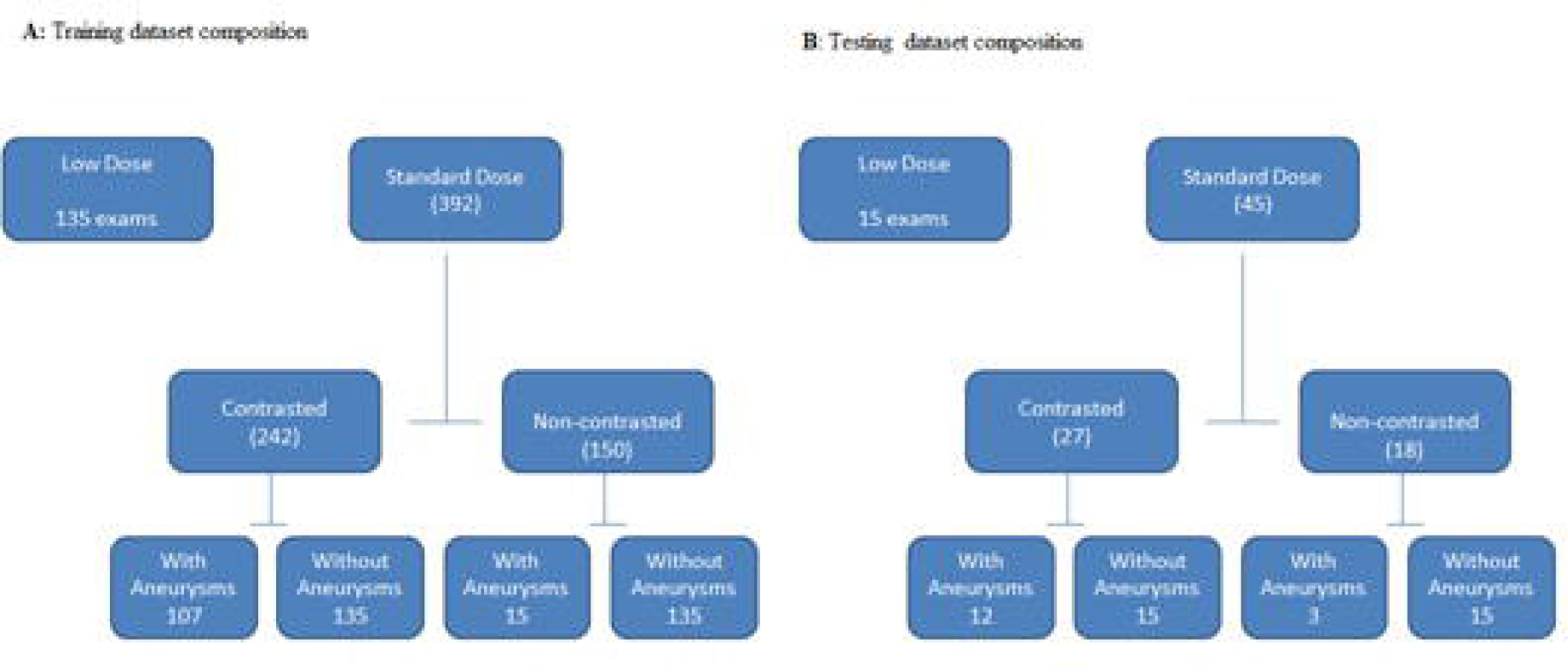
Dataset composition: A training dataset; B: validation dataset (Figure does **not** need to be coloured in print)

The original database was 796 exams, from which we excluded 210 based on our exclusion criteria. From the remaining 587 exams performed, 527 were used for data training and 60 for model validation (Figure 1).

Population characteristics are described in Table 1.

**Table 1.** Descriptive statistics.

|  |  | Low-Dose<br>(n = 135) | Std-Dose<br>(n=135) | CTA<br>(n=135) | Aneurysm<br>(n=107) | Aneurysm NC<br>(n=15) |
| --- | --- | --- | --- | --- | --- | --- |
|  |  | N (%) | N (%) | N (%) | N (%) | N (%) |
| **Patient** |  |  |  |  |  |  |
| **Position** | FFS | 150<br>(27.5%) | 130<br>(23.8%) | 137<br>(25.1%) | 113<br>(20.7%) | 16 (2.9%) |
|  | HFS | 0 (0%) | 20<br>(48.8%) | 13<br>(31.7%) | 6 (14.6%) | 2 (4.9%) |
| **Sex** |  |  |  |  |  |  |
|  | Male | 72<br>(20.6%) | 96<br>(27.4%) | 89<br>(25.4%) | 80<br>(22.9%) | 13 (3.7%) |
|  | Female | 78<br>(32.9%) | 54 (22.8) | 61 (25.7) | 39<br>(16.5%) | 5 (2.1%) |
| **Measurements** | **Average** | **Average** | **Average** | **Average** | **Average** | **Average** |
|  | **±Standard deviation** | **±Standard deviation** | **±Standard deviation** | **±Standard deviation** | **±Standard deviation** | **±Standard deviation** |
|  | Age | 60.93<br>±4.64 | 43.01<br>±18.37 | 55.89<br>±14.98 | 65.76<br>±12.96 | 69.72<br>±17.63 |
|  | Height | 1.67 ±0.09 | 1.71 ±0.09 | 1.70 ±0.10 | 1.70 ±0.09 | 1.73 ±0.9 |
|  | Weight | 73.60<br>±15.90 | 72.73<br>±16.92 | 74.82<br>±15.42 | 77.27<br>±17.70 | 83.71<br>±17.95 |

**Table 2.**
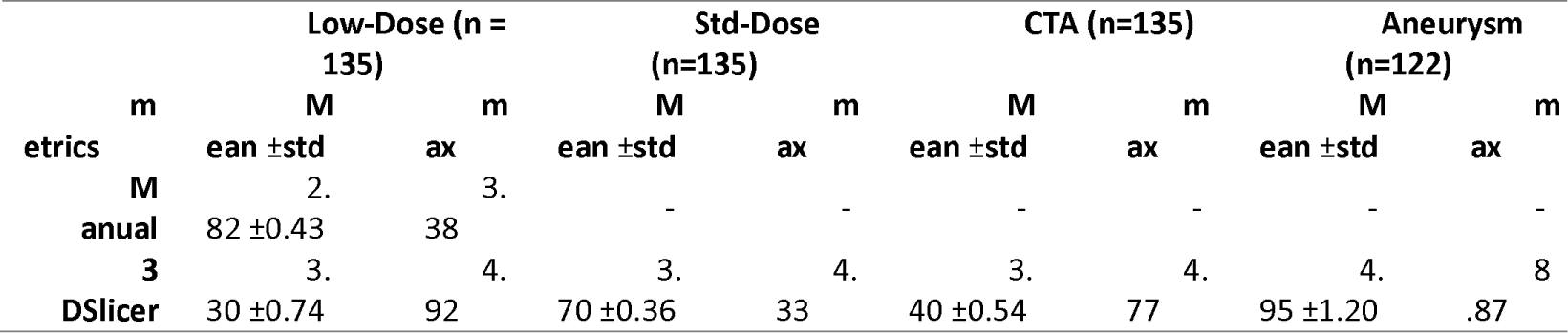
Comparison results of different methods of measuring the thoracic aorta.

## 3 Results

### 3.1 Manual segmentation and segmentation evaluation

A sample of 16 low-dose exams was selected to evaluate the marking of the aorta region in exams among physicians. The Dice Score Coefficient metric was used to assess markup quality (Supplementary Table 1).

The voting methodology was evaluated to unify the exam triplicates of this experiment. For this, two datasets were separated, a set with voting and another without voting. A DeepAAA model was trained with 50 epochs and compared the result of the average Dice Score Coefficient of these two datasets (Supplementary Table 2).

All exams were segmented, 527 masks, including triplicate segmented images submitted to the voting method.

### 3.2 Pre-processing

The processing of these exams was performed from the following steps, application of the adjustment of the conversion of the Hounsfield unit on each exam. In addition, windowing is applied to width 400, resampling of x-axis = 2mm, y-axis = 2mm, and z-axis = 3mm and cropping at 128×128×Z, where Z maintains its original exam value, per exam slice.

### 3.4 Training segmentation models

The average Dice Score for the DeepVox model, which required about 50 hours and 100 epochs of training, was 0.8708, while the average Dice Score for the DeepAAA model, which required about 113 hours and 100 epochs of training, was 0.87 (Supplementary Table 3).

### 3.5 Aorta diameter measuring

The largest measurement came from the “Aneurysm” dataset (8.87 cm) and the smallest from “Std-Dose” dataset (4.33 cm). Largest mean also came from “Aneurysm” dataset (4.95cm [std ±1.2]) but the smallest mean is from “Low-Dose” dataset (3.3 cm [std ±0.74]).

Table 3 displays the Maximum values and Mean Absolute Error (MAE) values achieved using automatic measuring technique and PyRadiomic.

**Table 3.**
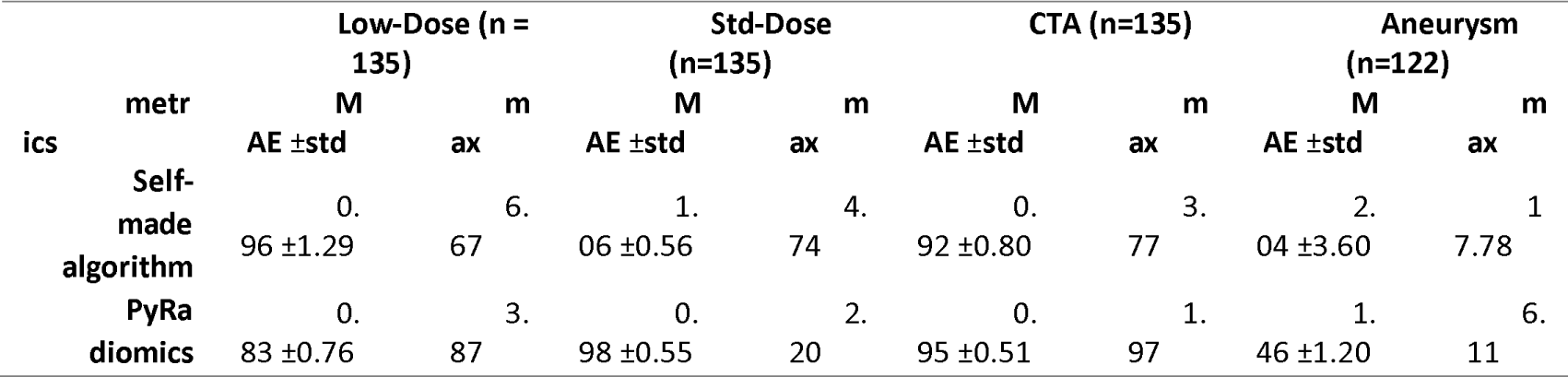
Comparison results of different methods of measuring the thoracic aorta.

The table shows that the PyRadiomics presented lower MAE values, all of which were less than 2mm. For the self-made algorithm approach, the absolute maximum values are greater than 10 mm, while for the PyRadiomics, they are less than 5 mm.

## 4 Discussion

In the present work we developed, applied, and evaluated a fully automated algorithm for the measurement of the thoracic aorta which delivered good results, when compared to manual aortic measurements.

Our Dice Score Coefficient results are very similar to results previously presented in the literature. Lu et al, 2019 exhibited a Dice Score Coefficient of 0.90 by training the DeepAAA model on 321 exams (223 unique patients), where 48% were contrast-enhanced and 77% containing abdominal aortic aneurysm, and validating on 57 unique exams, where 51% were contrast-enhanced and 51% contained abdominal aortic aneurysm. Comelli et al., 2020 performed a 5-fold cross-validation with 72 patients, using contrast-enhanced ECG-gated CT exams.

The measurements obtained by 3DSlicer are on average near manual measurement, while the self-made algorithm and PyRadiomics have little difference in how demonstrated by MAE value. This difference is similar to the results shown by Macruz et al^21^. But the maximum value by the self-made algorithm is showing must higher than PyRadiomiocs, we can infer that our measurement algorithm tends to underestimate the maximum diameter of the aorta. In addition, we consider that qualitatively the error of the measurement algorithm should be below 5 mm as shown in the literature.

From a clinical standpoint, our work addresses relevant issues. The development of an accurate automated measuring of the aorta, applicable even to low-radiation and non-contrasted exams, represents the possibility of increasing aneurysm diagnostic, which may, in turn, have a positive impact in mortality and rupture rates.

A MAE under 5mm, in clinical terms, is very accurate. The threshold for indicating surgical treatment in the thoracic aorta is usually over 4 to 6cm^1^, whereas clinical observation of aneurysms look for a yearly growth of over 1cm^1^.

Limitations to this tool are mostly related to fact that the aortic segmentation presents an imbalance issue, where the volume of interest is considerably smaller than the background. Although the Dice Score Coefficient is an established metric to assess the model performance, when the loss version (Dice loss, defined by 1-Dice Score Coefficient) is applied to class imbalanced problems, it often exhibits high precision, but low recall. Several approaches which aim to improve the imbalanced performance were evaluated, where the optimal function selected was the Hybrid Focal Loss (HFL)^22^ a combination of the Focal Tversky Loss and the Focal Loss.

These limitations notwithstanding, the results of our applied models indicate that the developed algorithm is able to accurately perform an automatic measurement of the thoracic aortic diameters in several exams automatically segmented, including sets acquired with low radiation dosage, the presence of aneurysms and non-contrasted exams.

Whilst current commercial solutions always require manual input, which introduces inter-operator variability, the tool proposed in this study delivers an objective, fully repeatable and systematic framework. The proposed solution also shortens the processing time, making it compatible with the clinical routine, and applicable to large series of exams for research purposes.

## 5 Conclusion

This study presents an effective automated solution for thoracic aortic measurement; the MAE values obtained for the measurement algorithm were under 5mm, with good results in sets of standard and low-radiation exams, as well as those acquired with or without contrast enhancement and those in which aneurysms were present. This solution thus presents a possibility for an auxiliary automation tool for the process of diameter measuring for the thoracic aorta, instrumental for diagnosis and management of several circulatory conditions.

## Data Availability

All data produced in the present study are available upon reasonable request to the authors.

## Supplementary material

**Supplementary Table 1:** Dice Score Coefficient comparison between doctors

|  | 1 vs 2 | 1 vs 3 | 2 vs 3 | Average |
| --- | --- | --- | --- | --- |
| Dice | 0.88 | 0.83 | 0.80 | 0.84 |

**Supplementary Table 2:** Voting experiment

| Scenario | with voting | without voting |
| --- | --- | --- |
| Avg. Dice | 0.88 | 0.81 |

**Supplementary Table 3:** Models comparison of cross-validation results. All models trained with the whole dataset.

|  | DeepAAA |  | DeepVox |  |
| --- | --- | --- | --- | --- |
|  | Dice Score Coefficient(test dataset) | Time per Epoch (min) | Dice Score Coefficient (test dataset) | Time per Epoch (min) |
| Fold 1 | 0.89 | 17.00 | 0.88 | 7.5 |
| Fold 2 | 0.87 |  | 0.88 |  |
| Fold 3 | 0.86 |  | 0.86 |  |
| Fold 4 | 0.85 |  | 0.86 |  |
| Average | 0.87 |  | 0.87 |  |

### Legend for Supplementary Figures

**Supplementary Figure 2:** Algorythm coding for section plane diameter calculation

(Figure does **not** need to be coloured in print)

