## Supplementary figures and images for "Artificial intelligence for automated thoracic aorta diameter measurement using different computed tomography protocols"

### Supplementary Figure 1

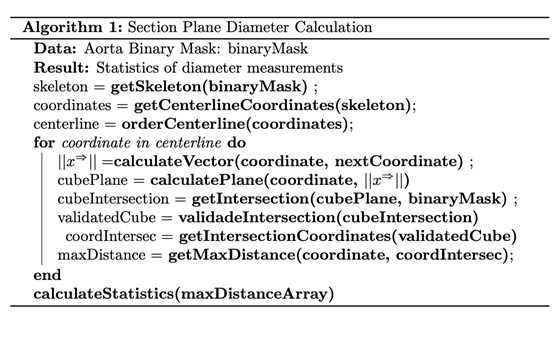
